# Prevalence and Associated Risk Factors of Scabies and Impetigo: A Cross-Sectional Study in the Nata Catchment Areas of Tutume District, Botswana

**DOI:** 10.1101/2023.07.08.23292236

**Authors:** Leungo Audrey Nthibo, Tuduetso Leka Molefi, Sidney Otladisa Kololo, Tshepo Botho Leeme, Mpho Selemogo, Mooketsi Molefi

## Abstract

**Background:** Scabies is a poorly understood disease in the developing world, particularly in regions with high disease burden. Lack of epidemiological data, especially in sub-Saharan Africa, hampers our understanding of the disease’s occurrence and impact. This study aimed to estimate the prevalence and associated risk factors of scabies and impetigo in the Nata catchment areas of Tutume district.

**Methodology:** A cross-sectional study was conducted in the Nata catchment area, targeting the settlements of Manxhotae, Malelejwe, Ndutshaa, and Tshwaane. Participants were randomly selected from randomly selected households. Data were collected using questionnaires, and scabies was confirmed by skin examination using the International Alliance for the Control of Scabies (IACS) consensus criteria. Statistical significance was set at p<0.05, with a 95% confidence interval for precision.

**Results:** A total of 429 participants were enrolled across the four settlements. The highest prevalence of scabies was in Manxhotae at 27.1% (21.2-34.0) and Ndutshaa at 23.4% (13.4-37.3). Malelejwe and Tshwaane had lower prevalence of 10.4% (6.2-16.8) and 3.4% (0.8-12.7), respectively. Only five(5) cases of impetigo were identified. Multivariable logistic regression analysis using the IACS criteria revealed that younger age and a household member with an itch were strongly associated with scabies, with adjusted odds ratios (AOR) of 5.69 [3.16-10.26] and 5.31 [2.76-10.21], respectively, however, a less sensitive criterion also included less frequent bathing as a significant exposure, AOR of 3.3 [1.9-5.8]

**Conclusion:** The prevalence of scabies in the Nata catchment area was unexpectedly high. The risk factors included younger age, a household member with an itch, and less frequent bathing. Prospective studies are needed to explore household disease transmission dynamics and risk factors specific to the youth.

**Author Summary:** This study was carried out in the Nata catchment area in Tutume district, Botswana. It aimed to assess the burden, risk factors informing effective disease control programs. The survey involved interviews and skin examination by a health worker. Findings revealed a higher incidence of scabies cases, while impetigo cases were less prevalent. These results highlight the need for community-wide interventions to mitigate the disease’s impact. Identified risk factors include younger age, residing with an individual experiencing itchiness, and infrequent bathing. Overall, this study supports advocating for scabies as a neglected tropical disease.

## Introduction

Scabies was added to the list of Neglected Tropical Diseases (NTDs) in 2017 due to its high burden and complications, particularly in areas with limited healthcare access(1). It is caused by a parasitic mite and often leads to impetigo, a bacterial skin infection. Lack of epidemiological data, especially in sub-Saharan Africa, hampers our understanding of the disease’s occurrence and impact. Scabies primarily spreads through direct skin-to-skin contact, and common risk factors include children, the elderly, immunocompromised individuals, overcrowding, and poverty(1-4). Scabies control efforts by the World Health Organization involve mapping disease burden, delivering interventions, and establishing monitoring and evaluation frameworks(1, 5). However, the lack of a field-friendly diagnostic test makes assessing scabies prevalence challenging.

In Botswana, as in many sub-Saharan African settings, the prevalence of scabies is poorly understood, with sporadic cases reported. Local epidemiological studies are needed to improve our understanding of the disease burden and associated risk factors, enabling the development of effective control programs. Previous experiences with other NTDs highlight the importance of mapping to successfully scale up control efforts(1, 2). This study aims to estimate the prevalence of scabies and impetigo, as well as identify associated risk factors in the Nata catchment area of Tutume district, Botswana. Such information will contribute to the implementation of appropriate and efficient disease control strategies.

## Methods

### Study design

This was a cross-sectional study whose main aim was to estimate the burden of scabies in the Nata catchment area following a confirmed outbreak. Analytic methods were used to determine possible risk factors associated with scabies.

### Study setting and participants

the study was conducted in the Nata catchment area, in Tutume district North-east of Botswana. Nata catchment area is predominantly a rural area. The population consists mainly of subsistence farmers and livestock herders. Health care services are provided through a Clinic in Nata, Health post in Manxhotae and mobile stops in the other settlements. Inclusion criteria included participants living in the selected communities, attending the selected schools and willing to participate in the survey. Individuals who did not live in the selected communities, who were unwilling to participate in the study and who were absent at the time of the visit were excluded from the study.

### Sampling

The study was performed during a mass drug administration campaign in the district following a confirmed scabies outbreak. Systematic sampling of households was done wherein study participants were randomly selected. In each household we enrolled every second person available.

Sample size calculation :the number of participants to be enrolled per site was determined using Cochran’s formula for sample size. Assuming a prevalence of 10% for the community, 7.5% margin of error and a design effect of 2.0 to cater for any clustering that may occur. For each of the sites, the following sample sizes were derived: Settlement Population Sample size Manxhotae 98 Malelejwe 94 Tshwaane 92 Nduutsha 88

### Data collection

prior to data collection community mobilization and sensitization was conducted by district healthcare workers in the communities that were to be surveyed. 1. Trained health care workers carried out a standardized skin examination in a private space set aside in the home/house. A skin examination of exposed regions of the body was performed on all participants. Scabies was diagnosed based on the criteria of the International Alliance for the Control of Scabies (IACS). (Appendix 1) Demographic information of the participants as well as clinical information was captured using a REDCap based questionnaire that was adapted from “A scabies outbreak in the North East Region of Ghana: The necessity for prompt intervention “ by Amoako and others.(6)

### Ethical considerations

written permission was obtained from the district health authorities. All participants were required to provide written informed consent. For children within the communities, written consent was obtained from parents or legal guardians and verbal assent was obtained from the children who wished to participate in the survey. All participants diagnosed with scabies or scabies and impetigo were treated with topical permethrin cream alone or in combination with cloxacillin.

### Statistical analysis

Data captured on REDcap software was transferred to Stata 13 (7) for further management and analysis. Demographic (e.g. sex), and clinical data (e.g. presence of an itch, rash etc) were summarized using frequencies and percentages for categorical variables, while for continuous variables e.g. age, normality tests were run and data summarised appropriately. Prevalence was calculated as the proportion of participants that report an itch or rash consistent with scabies or impetigo out of the total sample. Overall and per site prevalence was estimated. A binary regression logistic regression model was used to determine risk factors associated with the development of scabies. Whether as per international alliance for the Control of Scabies (IACS) consensus criteria has scabies or not was the dependent variable coded (1/0) in Stata, respectively. Several independent factors were evaluated as possible risk factors, these included sharing of bed or under-garments, bathing routine, presence of pipe-borne water in the homesteads. Confounding and effect modification were investigated and adjusted for. All measurements are reported with 95% precision and p-value of <0.05 is considered statistically significant.

## Results

A total of 429 participants were surveyed in all the selected villages, 188 (43.82%) in Manxotae, 135 (31.47%) in Malelejwe, 59 (13.75%) in Tshwaane, and 47 (10.96%) in Ndutshaa. The characteristics of the participants are displayed in table 1. 58% of the participants were female, while 41.75% were male. The median age is 35 years with an interquartile range of 25 and 56 years.

**Table 1:** Characteristics of study participants and proportion per village

| Characteristics | n (percentage) |
| --- | --- |
| Gender | (n=412) |
| Male | 172(41.75) |
| Female | 240(58.25) |
| Age | (n=380) |
| 0-9 | 39(10.26) |
| 10-19 | 60(15.79) |
| 20-29 | 73(19.21) |
| 30-39 | 76(20.00) |
| 40-49 | 36(9.47) |
| 50-59 | 28(7.37) |
| 60-65 | 19(5.00) |
| >65 | 49(12.89) |
| Village | (n=429) |
| • Manxhotae | 188(43.82) |
| • Malelejwe | 135(31.47) |
| • Tshwaane | 59(13.75) |
| Ndutshaa | 47(10.96) |

Table 2 shows prevalence of scabies by examination findings stratified per village. The village with the highest prevalence of scabies was Manxhotae, with a prevalence of 27.13%(21.22-33.96), followed by Nduutsha with a prevalence of 23.40% (13.35-37.73). Malelejwe and Tshwaane had a prevalence of 10.37% (6.22-16.80) and 3.39(0.84-12.74%) respectively.

**Table 2:** Prevalence of scabies by village using exam findings.

| Village | # with scabies (exam) | Population accessed | Prevalence (95%CI) |
| --- | --- | --- | --- |
| Manxotae | 51 | 188 | 27.13(21.22-33.96) |
| Malelejwe | 14 | 135 | 10.37(6.22-16.80) |
| Tshwaane | 2 | 59 | 3.39(0.84-12.74) |
| Ndutshaa | 11 | 47 | 23.40 (13.35-37.73) |
| Overall | 78 | 429 | 18.18(14.80-22.14) |

Using self-reported itch as proxy for scabies (see table 3), we note that in the unadjusted logistic regression model, age group less than 18 years of age (school going-age), those who reported someone an itch in the household, shared bedding and under-garments, and bathed other than once daily had higher odds of reporting an itch than their counterparts, OR=3.97[1.91-5.13],OR=5.20[3.26-8.29],OR=1.63[1.05-2.51], OR=1.52[0.91-2.52] and OR=2.31[1.42-3.74], respectively.

**Table 3:** Predictors of scabies using itch as proxy-unadjusted and adjusted binary logistic regression model

|  | Unadjusted analysis |  |  |  | Adjusted multivariable regression(n=363) |
| --- | --- | --- | --- | --- | --- |
| Variables | UO | SE± | p-value | 95% CI | AO 95% CI |
| **Demographics** |  |  |  |  |  |
| Age (years) (n=378) |  |  |  |  |  |
| 0-18 | 3.79 | 0.78 | 0.000* | 1.91-5.13 | 3.28 ( 1.87-5.76)* |
| >18 | 1 | - | - | - |  |
| Sex(n=410) |  |  |  |  |  |
| Female | 1.12 | 0.24 | 0.617 | 0.73-1.71 | - |
| Male | 1 | - | - | - |  |
| Number in household (n=411) | 0.99 | 0.03 | 0.944 | 0.94-1.06 |  |
| Exposures |  |  |  |  |  |
| Any who complained of the itch in the household (n=425) |  |  |  |  |  |
| Yes | 5.20 | 1.23 | 0.000* | 3.26-8.29 | 6.94 (3.92-12.29)* |
| No | 1 | - | - | - | - |
| Shared bedding (mattress, sheets etc) (n=425) |  |  |  |  |  |
| Yes | 1.63 | 0.36 | 0.030* | 1.05-2.51 | - |
| No | 1 | - | - | - | - |
| Shared room(n=398) |  |  |  |  |  |
| Yes | 1.08 | 0.08 | 0.328 | 0.93-1.26 | - |
| No |  |  |  |  |  |
| Shared under-garments(n=421) |  |  |  |  |  |
| Yes | 1.52 | 0.40 | 0.111* | 0.91-2.52 | 1.17 0.62-2.22 |
| No | 1 |  |  |  |  |
| Bath routine (n=416) |  |  |  |  |  |
| Other than daily | 2.31 | 0.57 | 0.001* | 1.42-3.74 | 2.06 1.42-4.80* |
| Daily | 1 | - | - | - |  |

\*-statistically significant 95% CI or p-value<0.05 unless otherwise stated

However, in the multivariable binary logistic regression model (see table 4) only younger age, those who reported to have had a household contact with an itch, and those who bathed other than daily had statistically significant odds of having scabies, OR=3.28 [1.87-5.76], OR=6.94[3.92-12.29] and OR=2.31[1.42-4.80]

**Table 4:** Predictors of scabies by examination of skin lesions-unadjusted and adjusted multivariable regression model

|  | Unadjusted analysis |  |  |  | Adjusted multivariable regression(n=363) |
| --- | --- | --- | --- | --- | --- |
| Variables | UO | SE± | p-value | 95% CI | AO 95% CI |
| Demographics |  |  |  |  |  |
| Age (years) (n=378) |  |  |  |  |  |
| 0-18 | 6.56 | 1.86 | 0.000* | 3.82-11.26 | 5.69 3.16-10.26* |
| >18 | 1 | - | - | - |  |
| Sex(n=410) |  |  |  |  |  |
| Female | 1.30 | 0.34 | 0.319 | 0.77-2.19 | - |
| Male | 1 | - | - | - |  |
| Number in household (n=411) |  |  |  |  |  |
|  | 1.68 | 0.43 | 0.040* | 1.01-2.78 | 0.98 0.55-1.75 |
| Exposures |  |  |  |  |  |
| Any who complained of the itch in the household (n=425) |  |  |  |  |  |
| Yes | 5.91 | 1.76 | 0.000* | 3.29-10.59 | 5.31 2.76-10.21* |
| No | 1 | - | - | - | - |
| Shared bedding<br>(mattress, sheets etc)<br>(n=426) |  |  |  |  |  |
| Yes | 2.25 | 0.363 | 0.004* | 1.30-3.90 | 1.10 0.59-2.06 |
| No | 1 | - | - | - | 1 |
| Shared room(n=399) |  |  |  |  |  |
| Yes | 0.99 | 0.64 | 0.991 | 0.28-3.55 | - |
| No | 1 | - | - | - |  |
| Shared under-garments(n=422) |  |  |  |  |  |
| Yes | 2.38 | 0.68 | 0.002* | 1.37-4.15 | 1.58 0.81-3.07 |
| No | 1 | - |  |  | 1 |
| Bath routine (n=417) |  |  |  |  |  |
| Other than daily | 1.41 | 0.41 | 0.236 | 0.80-2.51 | - - |
| Daily | 1 | - | - | - |  |

\*-statistically significant 95% CI or p-value<0.05 unless otherwise stated

In the diagnosis of scabies by examination of skin lesions, running an unadjusted regression model we noted that younger age (<18 years of age), increasing number of household members, those that reported someone else with an itch in the household, those who shared bedding and under-garments reported higher odds of scabies than their comparators, OR=6.56[3.82-11.26], OR=1.68[1.01-2.78], OR=5.91[3.29-10.59],OR=2.25[1.30-3.90] and OR=2.38[1.37-4.15], respectively however, in the multivariable binary logistic regression model only younger age and those who reported someone with an itch in the household emerged statistically significant, OR=5.69[3.16-10.26] and OR=5.31[2.76-10.21], respectively.

## Discussion

The burden of scabies was found to be highest in Manxhotae with a prevalence of 27.3% while overall prevalence for the Nata catchment area was found to be 18.8%. A systematic review which also included grey literature in all regions except North America found that the prevalence of scabies ranged from 0.2% to 71.4%, most regions had a prevalence more than 10%, this was in keeping with majority of population based studies in prevalence of scabies(2, 8-11). The high prevalence of scabies in the Nata catchment area which is like other studies suggests the need for intensified surveillance and case management to prevent further outbreaks. The high prevalence of the disease is suggestive of the unappreciation of scabies in resource-poor communities, and difficult access to the health system. The framework for scabies advises intensive clinical management of scabies for prevalence rates more than 10% and consideration for MDA(1). In most studies the prevalence of scabies in high burden countries decreased significantly after MDA (12-14). More than one cycle of MDA may be necessary to control the burden of the disease. Impetigo is found to be common, particularly in children. (3, 8, 15). Given that the study was performed during a confirmed scabies outbreak there may have been a degree of selection bias. The population on Nata catchment area also have a higher degree of poverty and overcrowding in households compared to the rest of the country. The villages selected for study are predominantly settlement type of villages. Other challenges also include shortage of water which may have played a role in the high prevalence. The International Alliance for the Control of scabies (IACS) has outlined a case definition for diagnosis of scabies however due to lack of objective diagnostic tests case under-ascertainment and underreporting may be a limiting factor in capturing the true picture of scabies cases on daily basis. Although scabies was more prevalent in the settlements, only a few cases of impetigo were identified. Five (5) cases of scabies and impetigo were identified, all from Manxhotae this was different from a population-based survey done in Fiji(10) which showed a prevalence of impetigo of 19.6% .other studies also showed a higher prevalence of co-infection of scabies and impetigo (16-18).The co-infection of scabies and impetigo is relatively common, especially in communities with poor hygiene and sanitation. This is because both conditions are highly contagious and tend to affect people living in close proximity to each other and crowded living conditions.

According to a study published in the Journal of Global Infectious Disease, the co-infection rate of scabies and impetigo in certain populations ranged from 10-83% and was particularly common in children (2). In some cases, the presence of scabies may increase the risk of developing impetigo because the intense itching caused by the mites can lead to skin damage and subsequent bacterial infection.

Analysing factors associated with scabies using itch as a proxy to scabies, in the multivariable logistic regression those who were <18 years, those who reported to have had someone with an itch in the household, and those who bathed other than daily had statistically significant odds of scabies. This finding is in keeping with most studies (8, 9). Nata catchment area is a settlement area with challenges such as poverty, shortage of water therefore the bathing routine is compromised resulting in most people bathing less frequently. Households have at most 5 people sharing a room which increases risk of transmission of human-to-human contact transmitted diseases. Sharing blankets and clothes is also a common practice which is culturally acceptable. This study did not show statistically significant results for association of higher household number with scabies which is different from other studies (6, 8, 17). Using examination findings to diagnose scabies in the unadjusted model younger age (<18 years of age); where the number of household members exceeds a median of 4; those that reported someone else with an itch in the household, and those who shared bedding and under-garments reported higher odds of scabies than their comparators. However, in the multivariable binary logistic regression model only younger age and those who reported someone with an itch in the household emerged statistically significant. Due to the weaker association between the other factors and scabies, the effect size diminished in the multivariable model. Although our findings are consistent with a systematic review and meta-analysis carried out in Ethiopia (11) which showed a correlation between the number of members per household and the risk of scabies, the magnitude of association found in our study was not as strong, and therefore the effect diminished in the multivariable model. (11)

This study possesses several strengths and limitations. It stands out as one of the few globally and regionally conducted studies shedding light on the neglected yet debilitating disease’s epidemiology. The analysis conducted in this study identifies established risk factors, such as a higher burden of the disease among the younger population, overcrowding, and its association with low socioeconomic status. However, the study did not explore other potential risk factors, such as the disease’s heightened risk among the HIV population.

Furthermore, while our multivariable regression models suggest potential associations between certain exposures and the occurrence of scabies, the lack of temporality between the exposure and outcome variables prevents us from fully establishing the findings. To provide a more comprehensive understanding of the associated factors, future epidemiological studies need to incorporate temporality, ensuring a clear delineation that exposure variables under investigation preceded the outcome.

## Conclusion

The study findings indicate a high overall prevalence of scabies and associated impetigo in the Nata Catchment area, with the highest prevalence observed in Manxhotae village. Our analysis of examination findings revealed that younger age and a history of someone with an itch in the household were identified as factors strongly associated with scabies.

## Data Availability

The data can be availed on request from the corresponding Author

## Acknowledgements

We would like to thank the Ministry of Health Botswana disease control division and the University of Botswana, department of Family Medicine & Public health leadership for affording the team the opportunity to carry out the investigation. We are equally thankful to the communities and participants who contributed to this study. Our gratitude is also extended to the Tutume DHMT community health nurse and health care assistants for helping with data collection and navigating the district.

## Data availability

Data is available on request to the corresponding author.

## Funding

There was no direct funding allocated to this study. Only Ministry of Health Botswana and University of Botswana resources were used.

## Author Contributions

**Conceptualization and Methodology:** Leungo Audrey Nthibo, Tuduetso Leka Molefi, Sidney Otladisa Kololo, Tshepo Botho Leeme, Mpho Selemogo, Mooketsi Molefi

**Investigation and Validation:** Leungo Audrey Nthibo, Tuduetso Leka Molefi, Sidney Otladisa Kololo, Tshepo Botho Leeme, Mooketsi Molefi.

**Formal analysis**: Mooketsi Molefi

**Supervision**: Mooketsi Molefi

**Writing – original draft**: Leungo Audrey Nthibo & Mooketsi Molefi

**Writing – review & editing**: Tuduetso Leka Molefi, Sidney Otladisa Kololo, Tshepo Botho Leeme, Mpho Selemogo

All authors read and approved the final manuscript.

**Corresponding author**

Correspondence to be addressed to Leungo Audrey Nthibo.

### Ethics declaration

The study was approved by the Ministry of Health human research and development Unit. Ref No:HRDP:6/13/1

### Conflict of interest

The authors declare that they have no conflict of interest.

## Notes

### Competing Interest Statement

The authors have declared no competing interest.

### Funding Statement

The author(s) received no specific funding for this work.

### Author Declarations

The Ethics committee of Republic of Botswana Ministry of Health under Health research division office gave ethical approval for this work

